# Clinical, Behavioral and Social Factors Associated with Racial Disparities in Hospitalized and Ambulatory COVID-19 Patients from an Integrated Health Care System in Georgia

**DOI:** 10.1101/2020.07.08.20148973

**Authors:** Felipe Lobelo, Alan Bienvenida, Serena Leung, Armand Mbanya, Elizabeth J Leslie, Kate E Koplan, S. Ryan Shin

## Abstract

**Introduction:** Racial and ethnic minorities have shouldered a disproportioned burden of coronavirus disease 2019 (COVID-19) infection to date in the US, but data on the various drivers of these disparities is limited.

**Objectives:** To describe the characteristics and outcomes of COVID-19 patients and explore factors associated with hospitalization risk by race.

**Methods:** Case series of 448 consecutive patients with confirmed COVID-19 seen at Kaiser Permanente Georgia (KPGA), an integrated health care system serving the Atlanta metropolitan area, from March 3 to May 12, 2020. KPGA members with laboratory-confirmed COVID-19. Multivariable analyses for hospitalization risk also included an additional 3489 persons under investigation (PUI) with suspected infection. COVID-19 treatment and outcomes, underlying comorbidities and quality of care management metrics, socio-demographic and other individual and community-level social determinants of health (SDOH) indicators.

**Results:** Of 448 COVID-19 positive members, 68,3% was non-Hispanic Black (n=306), 18% non-Hispanic White (n=81) and 13,7% Other race (n=61). Median age was 54 [IQR 43-63) years. Overall, 224 patients were hospitalized, median age 60 (50-69) years. Black race was a significant factor in the Confirmed + PUI, female and male models (ORs from 1.98 to 2.19). Obesity was associated with higher hospitalization odds in the confirmed, confirmed + PUI, Black and male models (ORs from 1.78 to 2.77). Chronic disease control metrics (diabetes, hypertension, hyperlipidemia) were associated with lower odds of hospitalization ranging from 48% to 35% in the confirmed + PUI and Black models. Self-reported physical inactivity was associated with 50% higher hospitalization odds in the Black and Female models. Residence in the Northeast region of Atlanta was associated with lower hospitalization odds in the Confirmed + PUI, White and female models (ORs from 0.22 to 0.64)

**Conclusions:** We found that non-Hispanic Black KPGA members had a disproportionately higher risk of infection and, after adjusting for covariates, twice the risk of hospitalization compared to other race groups. We found no significant differences in clinical outcomes or mortality across race/ethnicity groups. In addition to age, sex and comorbidity burden, pre-pandemic self-reported exercise, metrics on quality of care and control of underlying cardio-metabolic diseases, and location of residence in Atlanta were significantly associated with hospitalization risk by race groups. Beyond well-known physiologic and clinical factors, individual and community-level social indicators and health behaviors must be considered as interventions designed to reduce COVID-19 disparities and the systemic effects of racism are implemented.

## Introduction

As of July 2020, despite having 4.25% of the global population, the United States (US) has contributed a quarter of the more than 10 million cases and 500,000 deaths recorded globally due to coronavirus disease 2019 (COVID-19).(1) In the US — the current epicenter of the pandemic — it has been widely reported that Black/African Americans (AA) and other racial/ethnic minorities, particularly those living in large and diverse urban centers, shoulder a disproportionate burden of COVID-19 infection risk and associated adverse health outcomes.(2–6)

Earlier descriptive studies from patients admitted during March/April 2020 in Georgia showed an over-representation of COVID-19 hospitalizations and death rates among the Black/AA population.(7, 8) Subsequent reports from two large health care systems in Louisiana and California, and from the Veterans Affairs health system(9) also found racial disparities in COVID-19 outcomes and clinical risk factors for hospitalization. These reports also theorized that chronic disease control, health behaviors, social and other factors may contribute to such disparities.(3, 5),(7),(8) However, limited availability of quality of care history and social determinants of health metrics in most medical health records has precluded a comprehensive, broader analyses of potential drivers of these racial disparities.

The U.S. Census Bureau reports the racial/ethnic demographic distribution of Georgia as 58.3% White alone, 31.6% Black/AA, 9.7% Latino or Hispanic, and 4.1% Asian.(10) As of May 12^th^, the Georgia Department of Public Health (DPH) reported confirmed COVID-19 cases by race/ethnicity as 35.1% Black/AA, 30% white, 10.1% Hispanic or Latino and 1.4% Asian(11). This overrepresentation of Black/AA and other minority populations has been observed in other Georgia counties.(11) Kaiser Permanente Georgia (KPGA) is a regional integrated health care system serving over 300,000 members in 32 counties located in the Atlanta Metropolitan Area and the Northeast region of the state. As of April 2020, KPGA membership is 43% Black/AA, 30% White, 5% Asian, 4% Hispanic or Latino (18% Unknown/other), which more closely resembles metro Atlanta than overall Georgia.(12) In this study, we conducted a descriptive analysis of KPGA members with suspected and laboratory-confirmed COVID-19. KPGA’s robust electronic health record (EHR) data enabled analyses throughout the continuum of care including pre-pandemic underlying disease control, COVID-19 outpatient/inpatient management and post-discharge, with a particular focus on racial/ethnic comparisons. In addition, we conducted multivariable analyses for hospitalization risk based on demographics, comorbidities, quality of care metrics, lifestyle behaviors and other available individual and community-level social determinants of health indicators.

## Methods

For this retrospective cohort study, we performed an EHR review of KPGA members seen with COVID-19 related symptoms between March 3, 2020 and May 12, 2020. Patients were screened according to the Centers for Disease Control (CDC) and Georgia DPH guidelines.(13, 14) Patients who met criteria were tested for severe acute respiratory syndrome coronavirus 2 (SARS-CoV-2) by polymerase chain reaction (PCR). Due to limits on the testing capacity in Georgia during this period, patients with symptoms or exposures consistent with SARS-CoV-2 were categorized as having been tested or as a person under investigation (PUI). Patients who received tests were further categorized as confirmed or ruled-out. At the start of the epidemic in our region, KPGA prioritized testing among symptomatic health care workers and/or with relevant exposures and symptomatic KPGA members requiring hospital admission. After April 22, testing was progressively expanded to include clinical dispositions dependent on test results (pre-operative clearance, dialysis pending, skilled nurse facility or hospice placement), high risk symptomatic patients based on clinical criteria (>65 years, immunocompromise, chronic obstructive pulmonary disease (COPD), moderate-to-severe asthma, serious heart condition, Body Mass Index (BMI)>40, diabetes, chronic kidney disease (CKD), liver disease, pregnancy) and symptomatic patients with public health implications (non-KPGA healthcare workers, first responders, jail and elder care employees, etc.)

### Patient Demographics

We characterized confirmed SARS-CoV-2 and PUI patients by age, sex, self-reported race/ethnicity, insurance type, and area of residence. Race/ethnicity was categorized as non-Hispanic Black/AA (confirmed n=306; 68,3%), non-Hispanic White (n=81; 18%), and Other (n=61; 13,7%), which included Hispanic or Latino (n=16), Asian (n=15), Native American (n=1) and unknown/declined to report (n=29).

We obtained patient’s location of residence from the EHR and categorized it into four different regions of metro Atlanta: Northeast, Northwest, Southeast, and Southwest. Residence location was also linked to the neighborhood deprivation index (NDI), a composite SDOH measure including income, education, employment and housing quality.(15, 16) The higher the NDI value, the higher the level of deprivation in the neighborhood.(15, 16) We also utilized ESRI® Business Analyst data, a comprehensive demographic and lifestyle database which provides data to help interpolate patient’s socioeconomic status(17). Specifically, we linked patients’ places of residency with ESRI’s® zip code level classifications of median household income, occupation, and educational attainment. We used this data to cross-reference median household income with the government-defined poverty line.(18)

### Comorbidities

Existing comorbidities of SARS-CoV-2 confirmed patients were obtained from the patient’s EHR as classified by the International Statistical Classification of Diseases and Related Health Problems codes (ICD-10)(19). The Charlson Comorbidity Index (CCI) was used as a continuous measure of total comorbidity burden.(20) We used pharmacy dispensing data to compile the frequency of outpatient medications used by patients.

We used Healthcare Effectiveness Data and Information Set (HEDIS)(21) as a marker of hypertension (blood pressure reading lower than <140/90mm Hg) and diabetes control (glycated hemoglobin HbA1c < 8%) within a minimum rolling 12-month period.

Using KPGA’s Exercise Vital Sign (EVS) data, patient’s physical activity levels were classified as inactive, insufficiently active, and sufficiently active for those self-reporting ≤10 minutes, 11-149 minutes, and ≥150 minutes of exercise/week, respectively. The EVS has been previously validated(22) and is considered a clinically relevant screening tool for physical activity behaviors in the health care setting(23, 24).

### Clinical Outcomes

Hospitalized patients with confirmed COVID-19 were characterized by hospital length of stay (LOS), ICU LOS, invasive mechanical ventilation initiation and length of use, hospital discharge, readmission, currently hospitalized (n=20; 8.9%), and deceased. Instances of admission and discharge on the same date were defined as LOS of one day. Mechanical ventilation data was compiled using an ICD-10 code flagging instances of emergency endotracheal intubation during hospital stay. Once identified, a manual chart review was conducted for each eligible patient to calculate the length of mechanical ventilation, from intubation to extubation or death. Readmissions were defined as instances of subsequent admission within 30 days to a hospital after recent discharge. We conducted manual record reviews to distinguish between encounters of readmission and patient transfers from a hospital to another medical facility.

### Statistical Analysis

We report numbers (percentages) for binary and categorical variables and medians (interquartile ranges, IQR) for continuous variables. Chi-square tests ANOVAs and two sample t-tests were used to determine significant differences between groups. For two sample t-tests with statistically unequal variances, the Satterthwaite method was applied and reported.

Multivariable logistic regression was used to explore factors associated with having a COVID-19 related hospitalization in seven different models: COVID-19 confirmed cases only, confirmed cases plus PUIs, and confirmed cases plus PUIs stratified by race/ethnicity (Black/AA, White, Other) and by sex (Male and Female). All multivariable logistic regression models included age, gender and race/ethnicity as independent variables and hospitalization as the dependent variable. All additional independent variables were assessed using a bivariate analysis, either chi-squared or two sample t-test, and only the variables showing evidence of a statistically significant (α=0.05) relationship with the dependent variable were considered for entry into the models. A subset of the dependent variables was considered for the confirmed cases model due to the reduced sample size of the population. Stepwise selection method was used for final dependent variable selection with effect entry and effect remain significance levels of 0.05. All data analysis was conducted using SAS 9.4 software.

The KPGA institutional review board approved this study with a waiver of informed consent.

## Results

### Epidemiologic Characteristics

Within the study period we screened 6,568 patients, tested 2,920 (44.5%) and 448 (15.3% of tested) patients were positive for SARS-CoV-2. The median age of confirmed positive patients was 54 [IQR, 43-63] years old. Black/AA patients resided in neighborhoods with the highest rate under the federal poverty level (14.2%), unfavorable NDI (0.45), and the highest mean percentage of frontline (35.7%) and healthcare workers (7.5%). (Table 1). The highest percentage of the KPGA members with confirmed SARS-COV-2 resided in the Northeast Metro Atlanta area (31.5%). However, different areas of metro Atlanta showed varying prevalence of KPGA members with confirmed SARS-COV-2 when stratified by race/ethnicity. More Black/AA and Other race patients lived in the Southern areas of metro Atlanta which visible correlates with more socially deprived neighborhoods. (Figure)

**Table 1:**
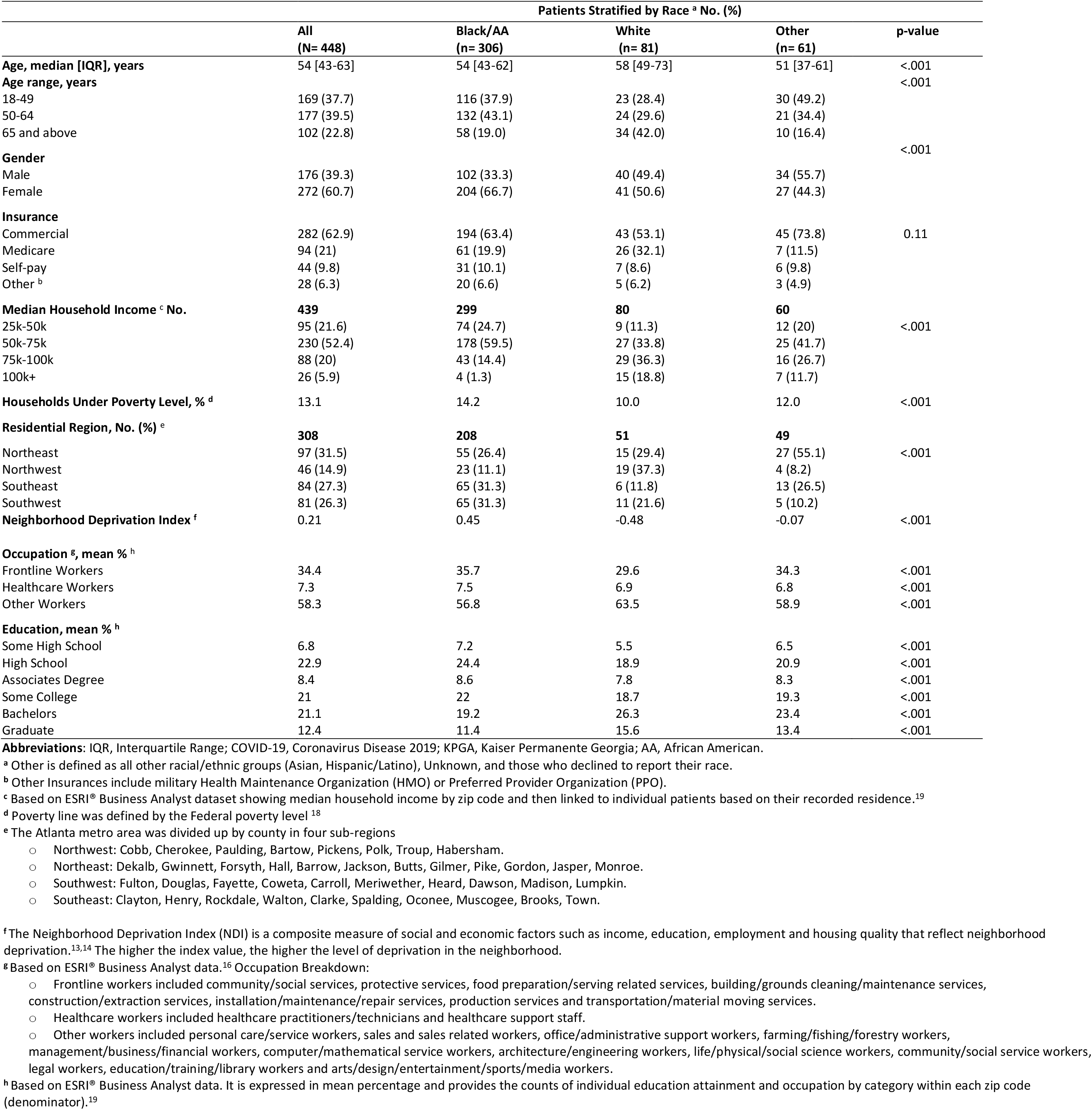
Socio-economic and demographic characteristics of KPGA members with confirmed SARS-COV-2 seen from March 3 to May 12, 2020.

**Figure 1.**
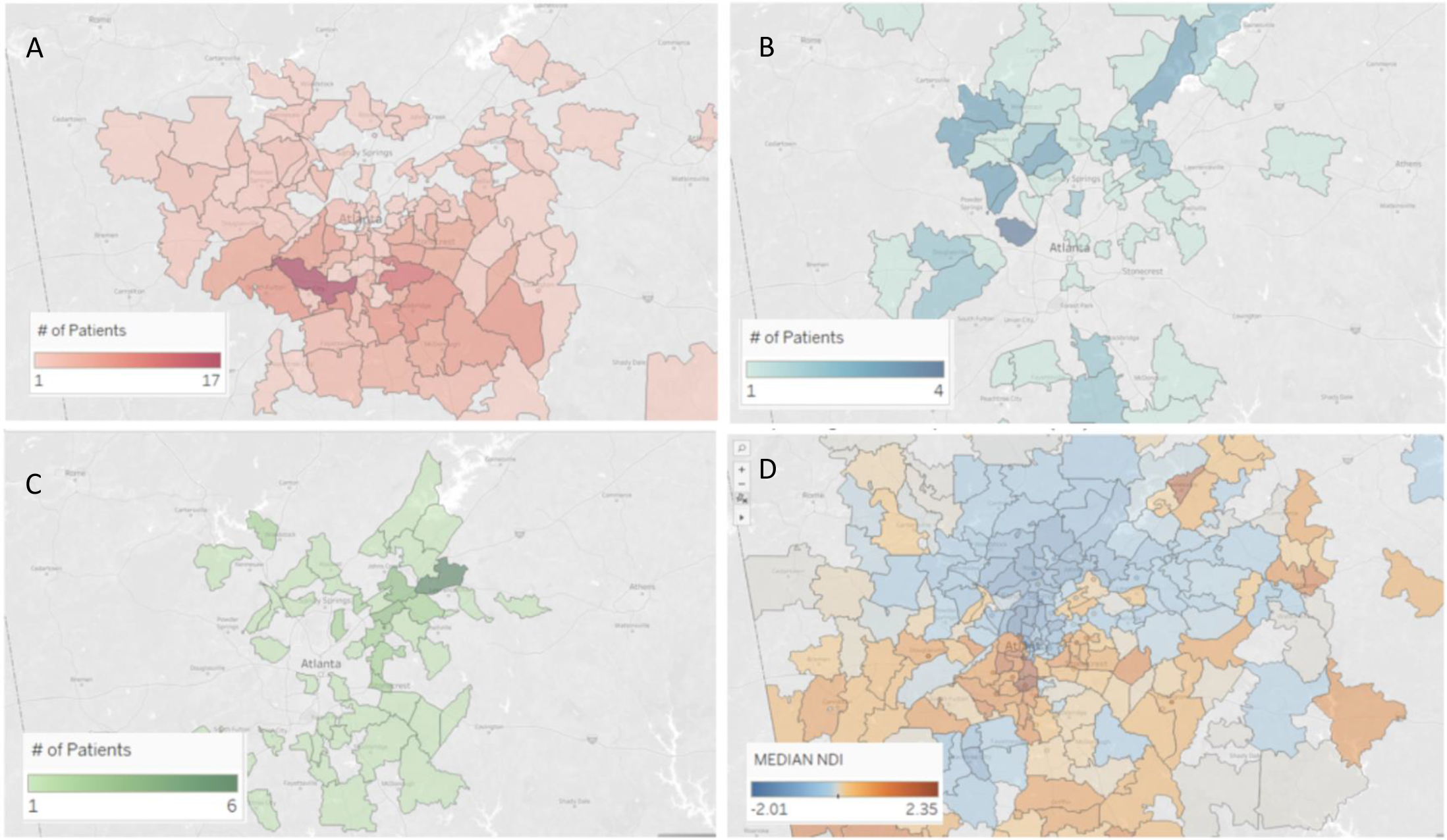
Heat Map of SARS-COV-2 Confirmed Positive Patients By Race/ethnicity Groups Across Metro Atlanta’s KPGA Catchment Area. Panel A: non-Hispanic Black/African American; Panel B: non-Hispanic White; Panel C: Other race group (Hispanic/Latino, Asian, American Indian, unknown/declined to report); Panel D: Metro Atlanta Neighborhood Deprivation Index

### Clinical Characteristics

Black/AA patients had higher rates of obesity (67.3%), hypertension (54.9%), and 2 or more comorbidities (66.3%). White patients presented with higher rates of hyperlipidemia (50.6%), congestive heart failure (CHF; 24.7%), coronary artery disease (CAD; 13.6%), arrhythmia (13.8%), CKD (11.1%) and overall CCI Scores (3.2 [2.2]) (all p<0.001) (Table 2).

**Table 2:**
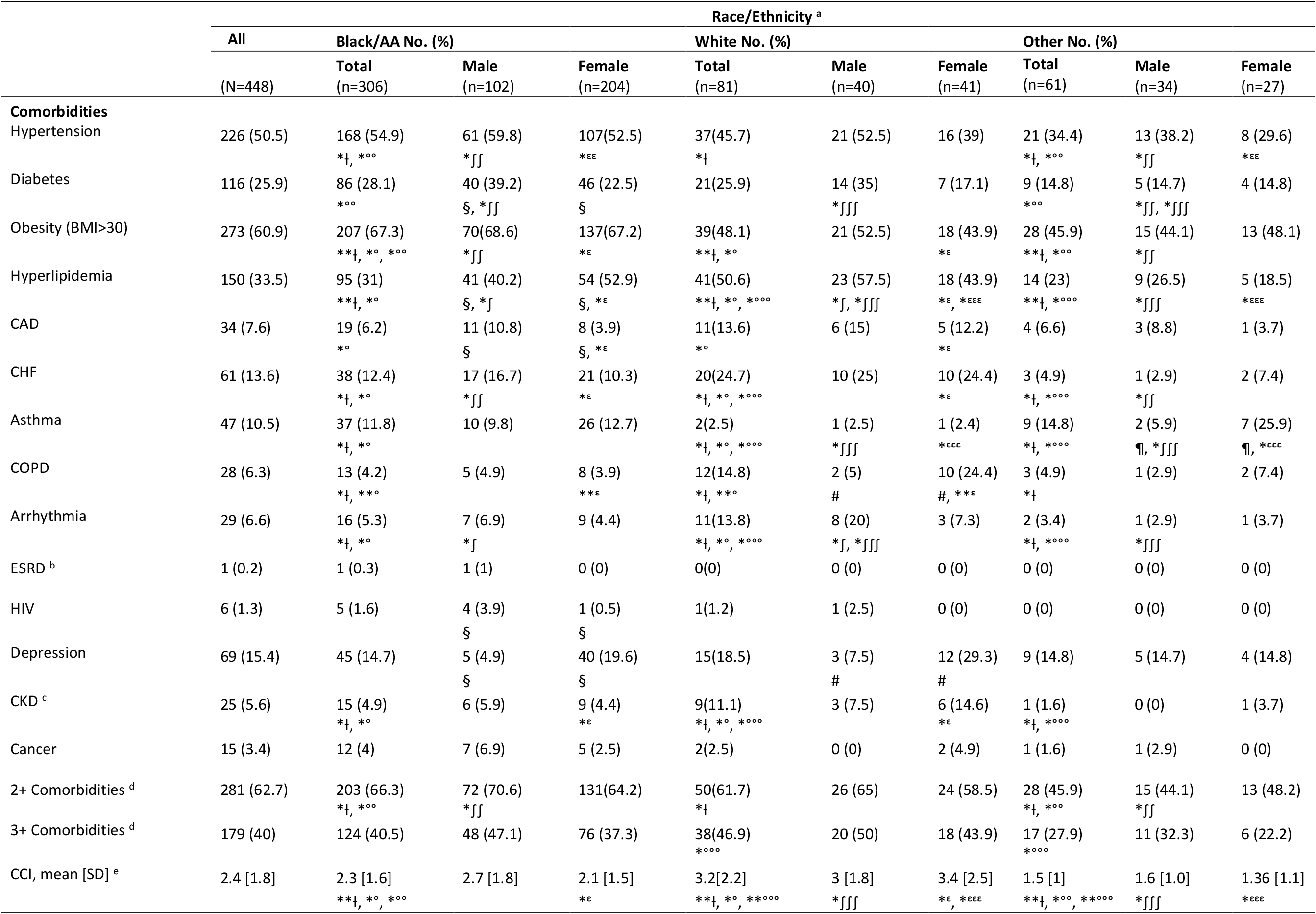
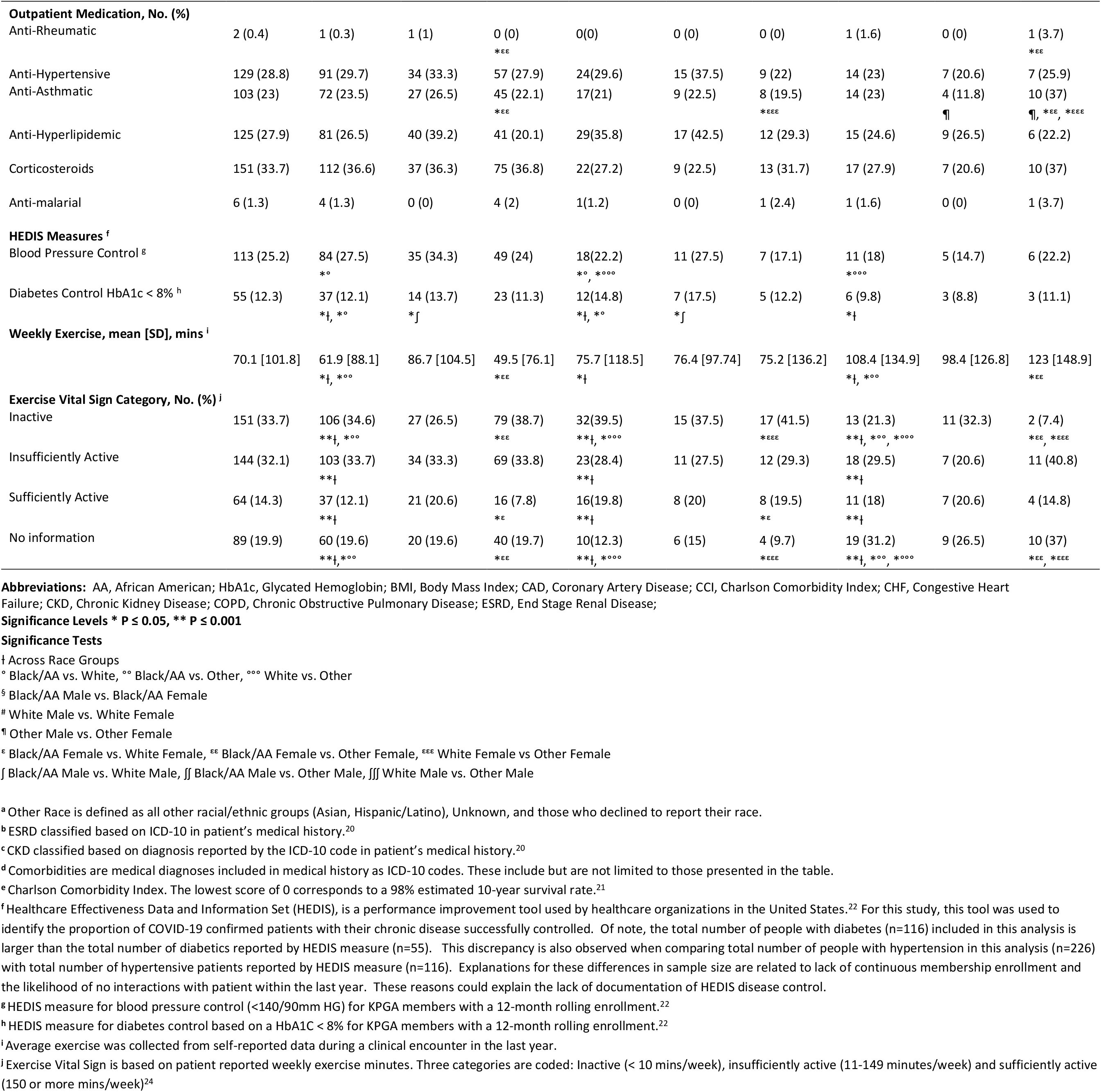
Comorbidities, outpatient medication, history of disease control, & exercise frequency of KPGA members with confirmed SARS-COV-2 by race & sex.

Compared to other race/ethnicity groups, White patients had the highest rate of diabetes control (14.8%) but Black/AA patients had higher rates of blood pressure control (27.5%) as measured by HEDIS measures. Black/AA patients self-reported the least mean [SD] average weekly exercise minutes in all race/ethnicity groups (61.9 [88.1]; p <0.001). The prevalence of physical inactivity was higher for both Black/AA and White females compared to Other race females (38.7% vs. 41.5% vs 7.4%; all p <0.001, respectively).

### Clinical Outcomes of Hospitalization

Overall, 224 patients with laboratory confirmed COVID-19 were hospitalized with 248 hospital stays, a median age of 60 (50-69) and a median length of stay of 6 (3-11.3) days. (Exhibit 3) There were no significant differences between Black/AA and White patients in ICU admission, ICU LOS, invasive mechanical ventilation and death (8.1% vs. 14.6%). Black/AA females were hospitalized on average 2.4 days longer than white females (95% CI 0.11 to 4.6; p ≤0.05). White females had higher 30-day readmission rates than Black/AA females (17.9% vs. 4%; p ≤0.05). Other race females showed significantly higher rates of invasive mechanical ventilation compared to Black/AA and White females (50% vs. 17% vs 10.7%, p ≤0.05; respectively) (Table 3).

**Table 3:**
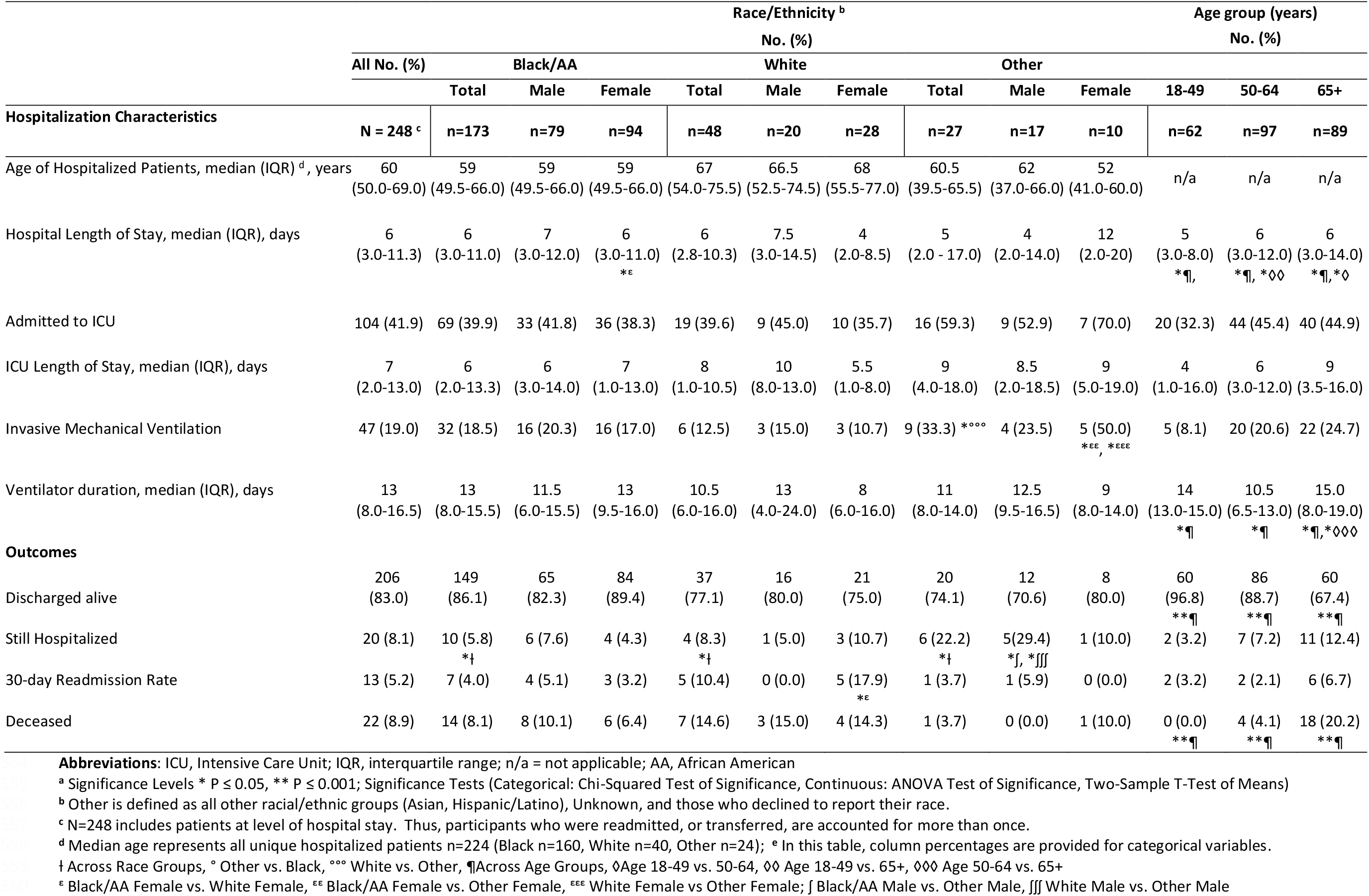
Clinical outcomes of hospitalized KPGA members with confirmed COVID-19 by race & age groups.

### Multivariable Analysis and Factors Associated with Hospitalization

Increasing age was a significant risk factor in all models and females had lower hospitalization odds in the confirmed, confirmed + PUI, Black/AA and Other race models (ORs ranging from 0.33 to 0.51) (Table 4). Black/AA race was a significant factor in the Confirmed + PUI, female and male models (ORs ranging from 1.98 to 2.19). Obesity was associated with higher hospitalization odds in the confirmed, confirmed +PUI, Black/AA and male models (ORs ranging from 1.78 to 2.77). Every point increase in the CCI Index showed increased hospitalization odds in the White model (OR 1.35 95% CI 1.15 to 1.59; p<0.001) while patients with 2 or more Comorbidities had higher hospitalization odds in the Female model (OR 2.38 95% CI 1.43 to 3.94; p<0.001). Cardio-metabolic disease management and control metrics (diabetes, hypertension, hyperlipidemia) were associated with lower odds of hospitalization ranging from 48% to 35% in the confirmed + PUI and Black/AA models.

**Table 4:** Multivariable logistic regression model odds ratios for hospitalization by COVID-19 status, Rrce, and sex.

| Sample Population | SARS-COV-2 Status |  | Race/Ethnicity Groups |  |  | Sex Groups |  |
| --- | --- | --- | --- | --- | --- | --- | --- |
|  | Confirmed | Confirmed & PUI | Black/AA | White | Other <sup>a</sup> | Female | Male |
| Total sample size | n=448 | n=3,937 | n=2,156 | n=981 | n=800 | n=2,536 | n=1,401 |
| Variables, OR (95% CI) |  |  |  |  |  |  |  |
| Age | 1.03**(1.02,1.05) | 1.05**(1.04,1.06) | 1.04**(1.03,1.06) | 1.05**(1.02,1.08) | 1.06**(1.03,1.09) | 1.04**(1.03,1.06) | 1.06**(1.05,1.08) |
| Race: Black |  | 1.98**(1.44,2.72) | n/a <sup>b</sup> | n/a <sup>b</sup> | n/a <sup>b</sup> | 2.19**(1.40,3.42) | 2.03*(1.31,3.13) |
| Female | 0.42**(0.27,0.65) | 0.51**(0.38,0.68) | 0.52**(0.37,0.73) |  | 0.33*(0.14,0.77) | n/a <sup>c</sup> | n/a <sup>c</sup> |
| Obesity (BMI ≥30) | 1.87*(1.19,2.93) | 1.80**(1.31,2.47) | 1.78*(1.21,2.64) |  |  |  | 2.77**(1.78,4.32) |
| CCI <sup>d</sup> |  |  |  | 1.35**(1.15,1.59) |  |  |  |
| CHF <sup>e</sup> |  | 1.83*(1.19,2.80) | 1.99*(1.20,3.31) |  |  | 2.65**(1.54,4.52) |  |
| Hypertension <sup>e</sup> | 2.09*(1.33,3.29) | 1.76*(1.21,2.55) | 2.02*(1.26,3.21) |  |  |  |  |
| CKD <sup>e</sup> | 11.04*(1.39,87.60) |  |  |  |  |  |  |
| Diabetes <sup>e</sup> |  | 1.76*(1.23,2.51) | 2.00*(1.27,3.13) |  |  |  |  |
| Physically Inactive <sup>f</sup> |  |  | 1.50*(1.05,2.13) |  |  | 1.49*(1.00,2.20) |  |
| COPD <sup>e</sup> | 5.37*(1.45,19.81) | 1.72*(1.02,2.90) |  |  |  |  |  |
| 2+ Comorbidities <sup>g</sup> |  |  |  |  |  | 2.38**(1.43,3.94) |  |
| Antihypertensive |  | 0.65*(0.46,0.94) |  |  |  |  |  |
| Antihyperlipidemic |  | 0.65*(0.45,0.95) |  |  |  |  |  |
| HEDIS <sup>h</sup> BP Control <sup>i</sup> |  |  | 0.62*(0.41,0.94) |  |  |  |  |
| HEDIS <sup>h</sup> Diabetes Control <sup>j</sup> |  |  | 0.52*(0.29,0.93) |  |  |  |  |
| NE County Residence <sup>k</sup> |  | 0.64*(0.47,0.89) |  | 0.22*(0.07,0.66) |  | 0.61*(0.38,0.95) |  |

**Abbreviations:** OR, Odds Ratio; CI, Confidence Interval; PUI, Persons Under Investigation; n/a, not applicable; BMI, Body Mass Index; AA, African American; CCI, Charlson Comorbidity Index; CHF, Congestive Heart Failure; CKD, Chronic Kidney Disease; COPD, Chronic Obstructive Pulmonary Disease; HEDIS, Healthcare Effectiveness Data and Information Set; BP: blood pressure
Significance Levels \* P ≤ 0.05, \*\* P ≤ 0.001

Self-reported physical inactivity was associated with 50% higher hospitalization odds in the Black/AA and Female models. Residence in the Northeast region of Atlanta was associated with lower hospitalization odds in the Confirmed + PUI, White and female models (ORs ranging from 0.22 to 0.64)

## Discussion

This study shows an over-representation of Black/AA populations and other minorities in both the outpatient and inpatient phases of care for COVID-19 in an integrated care system, similar to previous reports (4, 6–9) Although a higher number of Black/AA KPGA members with COVID-19 were hospitalized (69.8%) than other races, there were no significant differences between racial/ethnic groups in ICU admission and duration, invasive ventilation and duration, 30-day readmissions, and mortality. However, further stratification by sex showed Black/AA females were hospitalized on average 2.4 days longer than white females, white females had higher 30-day readmission rates than Black/AA females (17.9% vs. 4%; p ≤0.05) and Other race females had higher rates of invasive mechanical ventilation compared to both Black/AA and White females (50% vs. 17% vs 10.7%, p ≤0.05; respectively). Previous studies reported similar clinical outcomes between Black/AA and non-Black hospitalized COVID-19 patients in Georgia(7, 8) and some but not all previous report have also showed no difference in clinical outcomes between racial/ethnic groups.(9)

Compared to White KPGA members, a higher percentage of Black/AA members with COVID-19 were female, younger, and more likely to reside in neighborhoods with median household income less than $75,000. Furthermore, Black/AA patients also reside at a higher proportion in neighborhoods with the highest rate of households below the federal poverty level (14.2%), positive neighborhood deprivation index (0.45), and the highest percentage of frontline (35.7%) and healthcare workers (7.5%) compared to other racial groups. All of these factors were associated with an increased risk of COVID-19 infection in our findings. In addition, Black/AA patients had the highest prevalence of obesity, hypertension, and presence of 2 or more comorbidities, all associated with increased disease severity in our analysis, as was found in previous reports.(4) However, we found the comorbidity burden was somewhat different by race, with White patients in our sample being on average older and showing higher CCI scores compared to Black/AA patients, and a different mix of specific underlying conditions (hyperlipidemia, CAD, CHF, arrythmia, CKD). Although there is a high prevalence of obesity, diabetes and other chronic diseases in the US population,(25–27) these findings suggest that a different comorbidity profile may influence COVID-19 disease severity across racial groups.

Similar to previous reports, our multivariable analysis revealed males were consistently more likely to be hospitalized while increasing age, obesity and hypertension were predominant factors associated with higher odds of hospitalization in all models.(7, 8) Black/AA race, diabetes, COPD, CHF, and CKD were also significant factors in different models. Of note, in the confirmed + PUIs model we detected a significant protective effect of outpatient hypertension and lipid management. Furthermore, Black/AA patients with adequate HEDIS blood pressure and diabetes control were significantly less likely to be hospitalized. Overall, these findings confirm that although the presence of various comorbidities is associated with COVID-19 admission, emphasis on providing adequate clinical management of baseline cardio-metabolic diseases could help ameliorate hospitalization rates. As the pandemic progresses over time, enhanced measures to ensure high quality of care for patients with multiple comorbidities should be reinforced, including those that leverage novel avenues of care including telemedicine and patient-generated actionable data, as well as sustainable linkages with community resources.(28, 29)

Beyond demographic and underlying comorbidity burden and management, our analyses also took into account the potential role of additional SDOH, including indicators of education, economic stability, neighborhood and physical environment and lifestyle behaviors. Of these metrics, we found that residence in the Northeast area of metro Atlanta was one of the most powerful protective factors for hospitalization in the Confirmed + PUI model, particularly for White and female KPGA members. Counties in the NE region have consistently higher levels of safety, quality housing, green space, education and income and have a lower prevalence of obesity compared to the southern regions of KPGA’s catchment area(30, 31).

Furthermore, self-reported physical inactivity — engaging in less than 10 minutes of moderate to vigorous exercise/week — increased by 50% the odds of hospitalization among Black/AA and female populations. Several biologic mechanisms may explain this novel association. Physical inactivity is a consistent risk factor for a plethora of chronic diseases shown to also increase COVID-19 severity.(32) Increased inactivity and sedentary time and related comorbidities are also associated with an increased low-grade chronic inflammatory state,(33) which may contribute to the known increased systemic inflammatory effects of SARS-COV-2. In addition to being a modulator of inflammation, regular moderate exercise is also an important immunomodulator, particularly of the virus-fighting cytotoxic immune response.(34) This is reinforced by epidemiologic studies showing a link between moderate-to-vigorous regular exercise and a lower risk of upper respiratory tract viral infections – including influenza and pneumonia – as well as improved vaccine responses.(35) Although previous reports have shown that self-reported exercise is a predictor of clinical outcomes(24), it is noteworthy that physical inactivity remained a significant correlate of hospitalization risk in our study population, after adjusting for traditional “hard” risk factors such as age, BMI, comorbidity burden and therapeutic management. This reinforces the clinical value of promoting fitness and an active lifestyle, preferably outdoors, to reduce the risk of infection and disease severity of a novel infectious agent such as SARS-COV-2.(36)

This study has some limitations. Limited testing availability in the early stages of the pandemic — globally and in Georgia — led to prioritizing those with the most symptomatic and severe disease requiring admission, as well as testing healthcare workers to prevent further nosocomial infection. For this reason, we included in our analysis not only laboratory-confirmed but also persons under investigation seeking care with COVID-like symptoms. However, we acknowledge that not all PUIs would necessarily have SARS-COV-2. The target population in this analysis included only KPGA members that by definition have insurance and ready access to health care services. However, our analysis showed a diverse socioeconomic background of KPGA members. Merging racial/ethnic groups with low sample sizes into a combined “Other” race category was necessary for statistical power reasons but limits the interpretation of findings for this group. Finally, despite having some SDOH indicators in our member’s EHR, we also included neighborhood level data to extrapolate additional SDOH metrics. Ongoing investigation of drivers in COVID-19 disparities will benefit from more individual level SDOH data. Despite these limitations, by integrating underlying chronic disease management history, outpatient information, hospitalization, clinical outcomes and post-discharge follow-up data, this study provides a comprehensive longitudinal assessment of COVID-19 patients in relation to racial/ethnic disparities.

To our knowledge, this investigation is the first COVID-19 retrospective cohort to include a multivariate analysis on multiple measures of SDOH and pre-pandemic comorbidity management. Our study suggests that, within our sample of KPGA members with ready access to insurance and high quality of care within an integrated health system, Black/AA members were still being disproportionately affected by COVID-19 risk of infection and hospitalization. However, we found no significant differences in clinical outcomes such as ICU length of stay or mortality across race/ethnicity groups. Location of residence, a proxy for the overall community context of our patients, appears to be a factor strongly associated with increased infection risk among Black/AA and other minorities. SDOH have also shown to contribute to a more unfavorable baseline health status and therefore, can indirectly impact COVID-19 risk of hospitalization and severity.(6) In addition to age, sex, location of residence and presence of comorbidities, pre-pandemic self-reported exercise levels and underlying cardio-metabolic disease control may also significantly impact hospitalization risk in different race groups. Therefore, beyond well-known physiologic and clinical factors, individual and community-level social factors and health behaviors must be considered by clinicians, health care systems(37) and public health stakeholders (6) as interventions designed to reduce COVID-19 disparities and the systemic effects of racism(38) are implemented.

## Data Availability

Yes

## Acknowledgements

Special thanks to all the clinicians, providers and staff of the Southeast Permanente Medical Group and Kaiser Permanente Georgia.

